# Biologic Drug Survival in Psoriasis: A Systematic Review & Comparative Meta-Analysis

**DOI:** 10.1101/2020.07.13.20151340

**Authors:** A. I. Mourad, R. Gniadecki

## Abstract

**Background:** Drug survival studies have been utilized to evaluate the real-world effectiveness of biologics used in psoriasis. However, the increasing volume of drug survival data suffers from large variability due to regional differences in drug availability, patient selection and biologic reimbursement.

**Objectives:** To conduct a meta-analysis of biologic drug survival to determine comparative effectiveness of the biologics in a real-world setting.

**Methods:** Studies reporting drug survival for biologic therapy in psoriasis were identified by a systematic literature search. Hazard ratio data for drug discontinuation were estimated directly from published Kaplan-Meier estimator curves at year 1, 2 and 5 of treatment and compared pairwise for the following biologics: ustekinumab, adalimumab, etanercept, infliximab, secukinumab and ixekizumab. This pooled hazard ratios were used to estimate 2- and 5-year overall drug survival rates.

**Results:** Ustekinumab had the longest persistence at 2 years and 5 years among all biologics included in this meta-analysis. Adalimumab was superior to etanercept and infliximab at 5 years. Pooled 5-year drug survival rates for adalimumab, etanercept, and infliximab were 46.3%, 35.9% and 34.7%, respectively. 2- and 5-year data were not available for anti-IL-17 drugs, but at 1-year ustekinumab outperformed secukinumab, the latter being equal to anti-TNFs.

**Conclusions:** Ustekinumab is characterized by longer drug survival than TNF inhibitors and IL-17 inhibitors. Estimated pooled 2- and 5-year drug survival rates may serve as a useful tool for patient communication and clinical decision-making.

## INTRODUCTION

Psoriasis is a chronic, immune-mediated dermatologic disease mediated by three key cytokines: IL-23, TNF-α and IL-17.^1,2^ The therapeutic monoclonal antibodies targeting one of those three central cytokines, effectively suppress the disease short term but they gradually lose their efficacy long-term. Drug survival, sometimes referred to as the drug persistence, measures the time until treatment discontinuation and has been widely applied as a marker of the real-world therapeutic effectiveness of various biologic therapies in psoriasis.^3-6^ The persistence of the biologics in real world is positively associated with the efficacy and safety^7-9^ but it may also be influenced by factors unrelated to the efficacy of the drug, such as reimbursement policies or therapeutic guidelines. Limitations on the duration of reimbursement during the therapy cycle or forced switching to the cheapest biologics have been implemented in some European countries and may significantly affect drug survival. Therefore, the techniques that allow for data synthesis from a number of different registries would allow for a better assessment of the performance of the biologic in the real-world setting.

Drug persistence is usually presented using Kaplan-Meier curves and Cox logistic regression is used to determine drug half-life and the hazard ratios for drug discontinuation^6^ Individual patient data are usually not available for cumulative analyses and it has therefore been difficult to pool the Kaplan-Meier estimators to synthesize biologic drug survival in psoriasis from different centers.^4^ Here, we adopted the methodology developed by Tierney et al.^10^ to conduct a meta-analysis of hazard ratios reflecting drug discontinuation rates over a predefined period (1, 2 and 5 years). This enabled us to compare the drug survival of different biologics against each other as well as to determine pooled overall survival rates for each respective biologic, including best- and worst-case biologic survival rates, when the data was sufficiently available. We believe that our results provide an intuitively understandable measure of the chances of long-term drug efficacy in the real world setting for the patients and the professionals, and can be used in the therapeutic decision making.

## MATERIALS AND METHODS

The Preferred Reporting Items for Systematic Reviews and Meta-Analyses (PRISMA) was used to report the results of this study.^11^ The study is registered with PROSPERO (CRD42020162368).

### Search Strategy

A comprehensive literature search was conducted using the following electronic databases: Medline, Cochrane Library, EMBASE(Ovid), Scopus, and Web of Science. The search strategy focused on persistence of biologics in psoriasis including their respective synonyms (drug survival, treatment adherence). The search of the electronic database to identify eligible studies was performed on April 15, 2020. Medical subject heading terms (MeSH terms) were incorporated in the search strategy when applicable, which included ((“Medication Adherence”[Mesh]), “Psoriasis”[Mesh]), and “Biological Products”[Mesh])).

### Study Eligibility, Selection Criteria, and Screening

The authors screened abstracts of the articles for inclusion using the following inclusion criteria: study type (cohort or case-control trials), patient age ≥18 years, diagnosis of psoriasis with or without psoriatic arthritis, biologic therapy (adalimumab (ADA), etanercept (ETA), infliximab (INF), ustekinumab (UST), golimumab (GOL), ixekizumab (IXE), secukinumab (SEC), and guselkumab (GUS)). If there were drug survival data available for these biologics, then they were included in the current study. Studies were selected based on these inclusion criteria and the two independent reviewers selected the eligible articles via screening through titles and abstracts. Where applicable, abstracts, letters to the editors, and unpublished data were evaluated. The eligible articles that were included on the initial screening process were then further independently reviewed in an un-blinded fashion by the two reviewers by full-text review. Articles that did not fulfill the selection criteria above were excluded. The authors of the respective included studies were contacted, where applicable, to identify any additional studies or to obtain raw data if required.

### Data Extraction

One author (A.M.) extracted data from the articles included after full-text review. The data that were collected included the following: author, year of publication, study design, observation time, treatment periods, mean age, sex (% male), a summary of overall drug survival at 6 months, 1 year, 2 years, and 5 years (when applicable). The extracted data was reviewed for accuracy by the senior author (R.G.).

### Risk of Bias and Quality Assessment

Risk of bias and study quality was assessed by the two authors using the Quality in Prognosis Studies (QUIPS) tool.^12^ This tool is a validated evaluative tool for assessing risk of bias and includes the following six domains: study participation, study attrition, prognostic factors measurement, outcome assessment, study confounding, and statistical analysis/reporting. The prognostic factor of interest in the current study was the reported overall biologic drug survival for each respective biologic derived from Kaplan-Meier survival analysis. Each domain was evaluated based on the respective prompting questions as specified by the tool, and then given a total overall risk of bias score (L= low risk, M= moderate risk, H= high risk).^12^

### Statistical Analysis

Time-to-event data were extracted from the Kaplan-Meier estimators of overall drug survival using a graphical digitizer program (DigitizeIt, ver 1.6.2). Data was collected from the Kaplan-Meier estimator curves for potentially eligible biologics (ustekinumab, adalimumab, etanercept, infliximab, ixekizumab, secukinumab and guselkumab), if the data was sufficient enough for collection. The available data was incorporated into a comparative meta-analysis of hazard ratios using methodology designed by Tierney et al.^10^ This methodology allowed for the pair-wise comparisons of the above candidate biologics, which yielded pooled hazard ratios for each respective comparison (ie: pooled hazard ratio was estimated for all the available comparisons for UST vs. ADA) The hazard ratio data for the primary outcome were analyzed at two time points (2 years and 5 years). A comparative pooled meta-analysis of the above comparisons at 2 years and 5 years was conducted using a random-effects model (Review Manager, ver. 5.3). Estimated 2- and 5-year overall drug survival rates were calculated using the pooled hazard ratios and their corresponding ±95% confidence intervals (CI) were used to calculate best-case and worst-case drug survival rates. Secondarily, a meta-analysis of secukinumab vs. other biologics at 1-year was conducted.

## RESULTS

### Study Selection

A systematic literature search was completed and identified 798 studies after duplicates were removed. Of the articles that were screened, 48 were then screened via full-text review.

Evaluation of these articles in duplicate led to the inclusion of 29 cohort studies in the systematic review, and 29 studies were included in the meta-analysis (Figure S1). A summary of the individual overall drug survival rates that were already published for ustekinumab, adalimumab, etanercept, infliximab, secukinumab and ixekizumab was tabulated (Table S1). A table summarizing the pooled hazard ratios derived from the below analyses was made (Table 1).

**Table 1:**
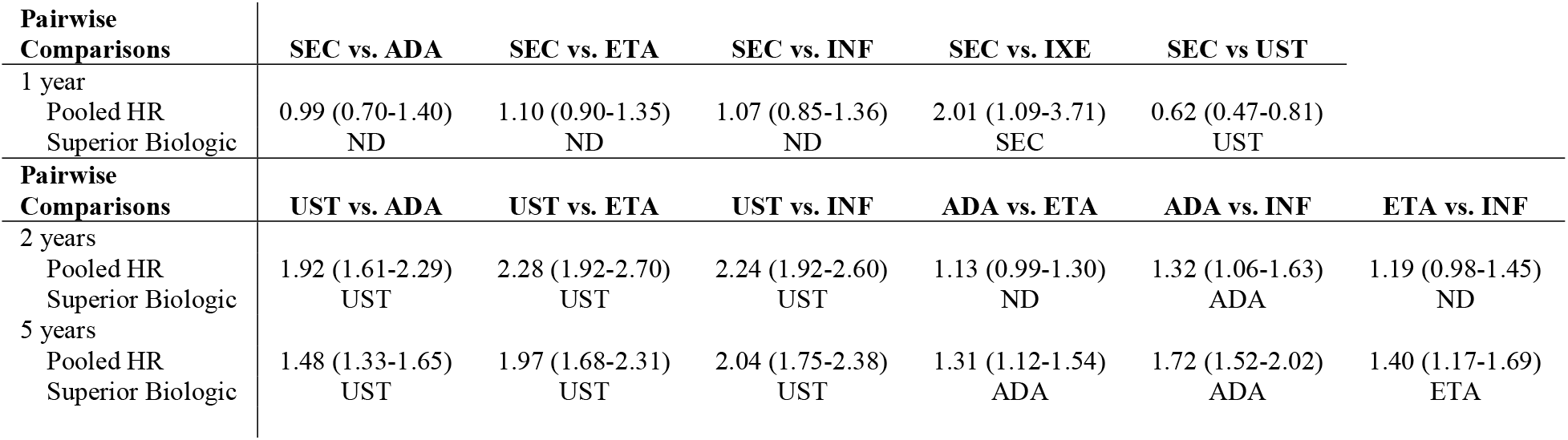
A summary of the pooled comparative analyses of biologic drug survival. Analyses were completed when there was sufficient data. UST: ustekinumab, ADA: adalimumab, INF: infliximab, HR: hazard ratio, ND: no difference, vs.: versus

| **Pairwise Comparisons** | **SEC vs. ADA** | **SEC vs. ETA** | **SEC vs. INF** | **SEC vs. IXE** | **SEC vs UST** |  |
| --- | --- | --- | --- | --- | --- | --- |
| 1 year<br>Pooled HR<br>Superior Biologic | 0.99 (0.70-1.40)<br>ND | 1.10 (0.90-1.35)<br>ND | 1.07 (0.85-1.36)<br>ND | 2.01 (1.09-3.71)<br>SEC | 0.62 (0.47-0.81)<br>UST |  |
| **Pairwise Comparisons** | **UST vs. ADA** | **UST vs. ETA** | **UST vs. INF** | **ADA vs. ETA** | **ADA vs. INF** | **ETA vs. INF** |
| 2 years<br>Pooled HR<br>Superior Biologic | 1.92 (1.61-2.29)<br>UST | 2.28 (1.92-2.70)<br>UST | 2.24 (1.92-2.60)<br>UST | 1.13 (0.99-1.30)<br>ND | 1.32 (1.06-1.63)<br>ADA | 1.19 (0.98-1.45)<br>ND |
| 5 years<br>Pooled HR<br>Superior Biologic | 1.48 (1.33-1.65)<br>UST | 1.97 (1.68-2.31)<br>UST | 2.04 (1.75-2.38)<br>UST | 1.31 (1.12-1.54)<br>ADA | 1.72 (1.52-2.02)<br>ADA | 1.40 (1.17-1.69)<br>ETA |

### Comparative Analyses

Comparative meta-analyses were conducted between ustekinumab, adalimumab, etanercept, infliximab to determine biologic which had the superior overall drug survival at 2 years and 5 years. 2- and 5-year biologic drug survival data was insufficient for secukinumab, ixekizumab, and guselkumab, and therefore meta-analyses at these time periods could not be conducted.

There were sufficient data for 1-year drug survival pairwise comparisons between secukinumab and the following biologics: adalimumab, etanercept, infliximab, ixekizumab, and ustekinumab. Therefore, the following pairwise comparisons of drug survival were completed as a secondary outcome at 1 year: SEC vs. ADA, SEC vs. ETA, SEC vs. INF, SEC vs. IXE, SEC vs. UST. Data for this secondary analysis is found below under “*Pooled Drug Survival at 1 year (secondary analysis*.

A meta-analysis of drug survival for ustekinumab vs. TNF-α inhibitors was conducted at 2 and 5 years (Figures S2, S3). The meta-analysis comparing drug survival for ustekinumab vs. adalimumab, etanercept, and infliximab at 2 years yielded pooled hazard ratios of 1.92 (95% CI: 1.61-2.29), 2.28 (95% CI: 1.92-2.70), and 2.24 (95% CI: 1.92-2.60), respectively (Figure S2). Pooled hazard ratios at 5 years were 1.48 (95% CI: 1.33-1.65), 1.97 (95% CI: 1.68-2.31) and 2.04 (95% CI: 1.75-2.38) for ustekinumab vs. adalimumab, etanercept, and infliximab, respectively (Figure S3). This data suggests the superiority of ustekinumab when compared against adalimumab, etanercept and infliximab. These data are summarized in Tables 1 and 2.

A comparative meta-analysis between the TNF-α inhibitors was completed at 2 and 5 years. There was no difference between the overall drug survival of adalimumab compared to etanercept at 2 years (HR: 1.13 (95% CI: 0.99-1.30)) (Figure S4). At 5 years, adalimumab was superior to etanercept (HR: 1.31 (95% CI: 1.12-1.54)) (Figure S5). Adalimumab had superior drug survival rates when compared to infliximab at 2 years (HR: 1.32 (95% CI: 1.06-1.63)) and 5 years (HR: 1.75 (95% CI: 1.52-2.02)) (Figures S4, S5). The pooled comparison between etanercept and infliximab revealed no significant difference between the two biologics at 2 years (HR: 1.19 (95% CI: 0.98-1.45)) (Figure S6). At 5 years, etanercept demonstrated superior drug survival when compared directly to infliximab (HR: 1.40 (95% CI: 1.17-1.69)) (Figure S7).

### Pooled Drug Survival at 1 year

Pooled analysis for secukinumab drug survival at 1 year revealed no statistically significant difference between secukinumab and the TNF-α inhibitors (Figure S8). Secukinumab showed superior pooled drug survival when compared to ixekizumab (HR: 2.01 (95% CI: 1.09-3.71)), although ustekinumab did show superior drug survival when compared to the same (HR: 0.60 (95% CI: 0.47-0.81)) (Figure S8).

### TNF-α inhibitor overall drug survival rates at 2 and 5 years

Pooled overall 2 and 5-year drug survival rates were calculated for adalimumab, etanercept, and infliximab. The pooled drug survival rate of adalimumab at 2 years was 53.2%, and 46.3% at 5 years (Figures 1a, 1b). The pooled drug survival rate for etanercept at 2 years was 47.6%, and 35.9% at 5 years. The pooled drug survival rate for infliximab at 2 years was 48.9%, and 34.7% at 5 years. We also calculated the survival percentages for 2 and 5 years in the best- and worst-case scenarios (i.e. for patients who are in the top or the bottom 5% of responders) (Figures 1a, 1b).

**Figure 1:**
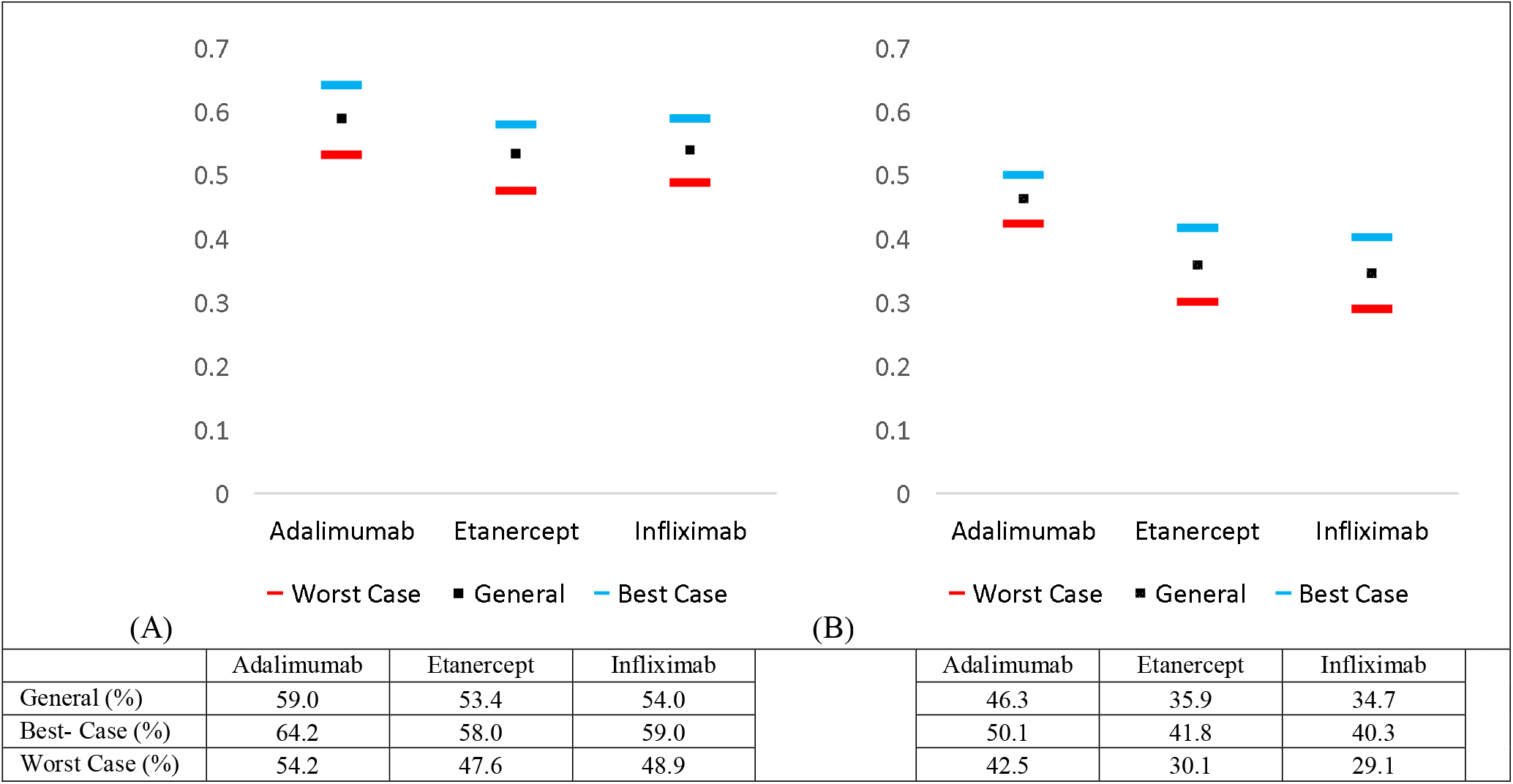
Estimated pooled 5-year drug survival rates for adalimumab, etanercept, and infliximab including worst- and best-case drug survival rates at (A) 2 years and (B) 5 years.

## DISCUSSION

The current study compares the real-life drug survival of biologics via meta-analysis of hazard ratios. The IL-12/23 inhibitor (ustekinumab) demonstrated superior biologic persistence when compared to TNF-α inhibitors (adalimumab, etanercept and infliximab). The analysis comparing the TNF-α inhibitors revealed that adalimumab was superior to etanercept and infliximab at year 5. Of note, there was no difference between the pooled drug survivals for adalimumab vs. etanercept and etanercept vs. infliximab at year 2. The data suggests that adalimumab is better suited for long-term adherence given its stronger performance at year 5 relative to etanercept and infliximab (Table 1). Pooled best- and worst-case drug survival rates were also calculated at 2 and 5 years (Figures 1a, 1b).

Strengths of this study include the utilization of robust statistical methodology to estimate hazard ratio data directly from the published Kaplan-Meier estimator data in the included studies. As biologic drug survival serves as a real-world surrogate for the efficacy and suitability for various biologic therapies used to treat psoriasis. Risk of bias, as determined by the QUIPS tool was low for the included studies. The pooled meta-analysis of these hazard ratios revealed moderate to high heterogeneity, which could be explained by varying factors intrinsic to the studies themselves (differences in patient populations, dosages, and registries). Currently available data on drug survival for the newer biologics used to treat psoriasis (secukinumab, guselkumab, ixekizumab etc) were limited, and as such, were not eligible for the pooled analysis at 2 and 5 years. We did; however, attempt to ameliorate this limitation to some extent by conducting the pooled analysis at 1 year for secukinumab. This allowed for the pair-wise meta-analysis between secukinumab and other biologics (adalimumab, etanercept, infliximab, ixekizumab, and ustekinumab). At year 1, secukinumab was superior to ixekizumab; however, there was no difference when secukinumab was compared to the TNF-α inhibitors. Interestingly, ustekinumab drug survival was remained superior when compared to pooled secukinumab drug survival. Given the lack of data beyond 1 year, we underscore the importance of having more long-term studies for these newer biologics.

To our knowledge, this is the first meta-analysis which conducts a direct pair-wise comparative analysis of hazard ratios of biologics in psoriasis. The information brought forth by this meta-analysis is dually useful in the fact that it could be used to guide treatment decision-making. This data could also serve as a vital tool for communicating the suitability of the different biologics when clinicians engage in real-life therapeutic discussions with their patients.

## Data Availability

Statistical used to run the meta-analysis is published in Mendeley. Mourad, Ahmed; Gniadecki, Robert (2020), Biologic Drug Survival in Psoriasis: A Systematic Review & Comparative Meta-Analysis, Mendeley Data, V1, doi: 10.17632/jb2dz98yn7.1

http://dx.doi.org/10.17632/jb2dz98yn7.1

## ACKNOWLEDGEMENTS

The study was supported by the Canadian Institutes of Health Research (CIHR) and the Canadian Association of Psoriasis Patients in the form of a medical student research studentship.

## FIGURE LEGEND

**Figure S1:**
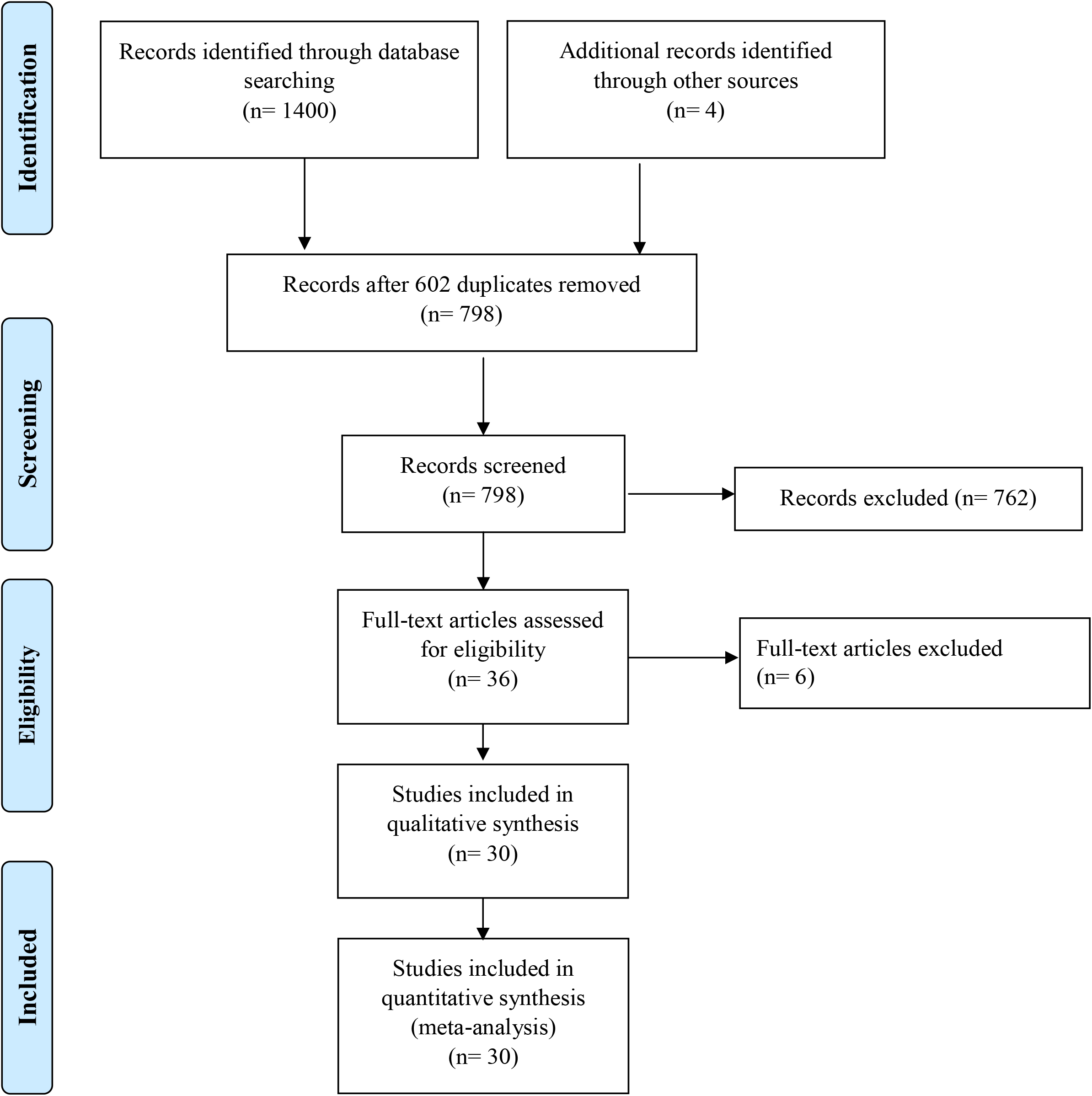
Flowchart of study selection process in accordance with PRISMA guidelines.

**Table S1:** Overall drug survival rates of ustekinumab, adalimumab, etanercept, infliximab, and secukinumab at 6 months, 1 year, 2 years, and 5 years. yr.: year.

|  | Ustekinumab |  |  | Adalimumab |  |  | Etanercept |  |  | Infliximab |  |  | Secukinumab |  |
| --- | --- | --- | --- | --- | --- | --- | --- | --- | --- | --- | --- | --- | --- | --- |
|  | 1 yr. | 2 yr. | 5 yr. | 1 yr. | 2 yr. | 5 yr. | 1 yr. | 2 yr. | 5 yr. | 1 yr. | 2 yr. | 5 yr. | 6mo. | 1yr. |
| **Arnold 2016<sup>1</sup>** | 0.90 | 0.83 | 0.75 | 0.70 | 0.53 | 0.49 | 0.60 | 0.48 | 0.29 | 0.54 | 0.37 | 0.11 | - | - |
| **Davila-Seijo 2016<sup>2</sup>** | 0.80 | 0.62 | 0.39 | 0.67 | 0.45 | 0.22 | 0.56 | 0.38 | 0.17 | 0.66 | 0.47 | 0.26 | - | - |
| **Egeberg 2016<sup>3</sup>** | 0.82 | 0.74 | 0.60 | 0.76 | 0.64 | 0.50 | 0.67 | 0.50 | 0.30 | 0.70 | 0.51 | 0.32 | 0.84 | 0.68 |
| **Esposito 2013<sup>4</sup>** | 0.93 | 0.84 | 0.64 | 0.88 | 0.69 | 0.45 | - | - | - | 0.80 | 0.60 | - | - | - |
| **Gniadecki 2011<sup>5</sup>** | - | - | - | 0.69 | 0.50 | - | 0.74 | 0.58 | - | 0.87 | 0.77 | - | - | - |
| **Gniadecki 2014<sup>6</sup>** | 0.83 | 0.74 | - | 0.75 | 0.65 | 0.50 | 0.70 | 0.55 | 0.34 | 0.72 | 0.61 | 0.42 | - | - |
| **Iskandar 2018<sup>7</sup>** | 0.85 | 0.77 | - | 0.74 | 0.58 | - | 0.49 | 0.36 | - | - | - | - | - | - |
| **Izinger 2016** | - | - | - | 0.71 | 0.57 | 0.48 | 0.71 | 0.60 | 0.44 | 0.58 | 0.30 | 0.12 | - | - |
| **Jacobi 2015<sup>8</sup>** | 0.90 | 0.75 | - | 0.70 | 0.54 | 0.44 | 0.74 | 0.49 | 0.24 | 0.51 | 0.17 | - | - | - |
| **Lunder 2018<sup>9</sup>** | 0.80 | 0.72 | - | 0.47 | 0.47 | - | - | - | - | 0.53 | 0.41 | 0.23 | 0.56 | - |
| **Marinas 2018<sup>10</sup>** | 0.91 | 0.86 | 0.76 | 0.80 | 0.65 | 0.51 | 0.83 | 0.69 | 0.42 | 0.84 | 0.66 | 0.38 | - | - |
| **Menter 2016<sup>11</sup>** | 0.95 | 0.90 | - | 0.75 | 0.64 | 0.41 | 0.67 | 0.58 | 0.49 | 0.83 | 0.78 | 0.53 | - | - |
| **Menting 2014<sup>12</sup>** | 0.85 | 0.68 | - | 0.84 | 0.75 | 0.57 | 0.86 | 0.69 | 0.27 | 0.68 | 0.52 | 0.43 | - | - |
| **Ohata 2018<sup>13</sup>** | 0.83 | 0.73 | 0.56 | 0.54 | 0.46 | 0.35 | - | - | - | 0.50 | 0.40 | 0.30 | - | - |
| **Pogacsas 2017<sup>14</sup>** | 0.86 | 0.75 | - | 0.69 | 0.58 | - | 0.72 | 0.60 | - | 0.71 | 0.50 | - | - | - |
| **Ross 2015<sup>15</sup>** | 0.91 | 0.91 | - | 0.73 | 0.53 | - | 0.57 | 0.48 | - | 0.68 | 0.68 | - | - | - |
| **Shalom 2017<sup>16</sup>** | 0.75 | 0.65 | - | 0.29 | 0.21 | - | 0.29 | 0.12 | - | 0.27 | - | - | - | - |
| **Sruamsiri 2018<sup>17</sup>** | 0.8 | 0.73 | 0.52 | 0.46 | 0.46 | - | - | - | - | 0.53 | 0.41 | 0.23 | 0.75 | 0.75 |
| **Verma 2018<sup>18</sup>** | 0.82 | 0.74 | 0.54 | 0.67 | 0.55 | 0.42 | 0.64 | 0.51 | 0.30 | 0.66 | 0.46 | 0.30 | - | - |
| **Vilarrasa 2016<sup>19</sup>** | 0.77 | 0.69 | - | 0.69 | 0.53 | - | 0.71 | 0.53 | - | 0.69 | 0.54 | - | - | - |
| **Warren 2015<sup>20</sup>** | 0.89 | 0.81 | - | 0.79 | 0.67 | - | 0.7 | 0.51 | - | 0.65 | 0.50 | - | - | - |
| **Zweegers 2016<sup>21</sup>** | 0.84 | 0.73 | - | 0.75 | 0.59 | 0.41 | 0.75 | 0.59 | 0.34 | - | - | - | - | - |
| **Cozzani 2019<sup>33</sup>** | 0.99 | 0.81 | 0.62 | 0.77 | 0.61 | 0.32 | 0.80 | 0.62 | 0.35 | 0.75 | 0.57 | 0.28 | 1.00 | 1.00 |
| **Kishimoto 2020<sup>34</sup>** | 0.86 | 0.79 | 0.56 | 0.71 | 0.63 | 0.52 | - | - | - | 0.58 | 0.50 | 0.31 | 0.85 | 0.68 |
| **Shalom 2020<sup>35</sup>** | 0.69 | 0.55 | 0.37 | 0.52 | 0.38 | 0.25 | 0.50 | 0.40 | 0.25 | 0.47 | 0.28 | 0.17 | 0.62 | 0.24 |
| **Egeberg 2018<sup>36</sup>** | 0.82 | 0.74 | 0.60 | 0.76 | 0.64 | 0.49 | 0.68 | 0.50 | 0.30 | 0.71 | 0.52 | 0.32 | 0.84 | 0.67 |
| **Potenza 2017<sup>37</sup>** | - | - | - | 0.84 | 0.74 | 0.65 | 0.81 | 0.72 | 0.53 | 0.71 | 0.51 | 0.36 | - | - |
| **Svedbom 2019<sup>38</sup>** | - | - | - | 0.68 | 0.55 | 0.31 | 0.62 | 0.45 | 0.15 | - | - | - | - | - |
| **Yiu 2020<sup>39</sup>** | 0.90 | 0.83 | - | 0.82 | 0.71 | - | - | - | - | - | - | - | 0.98 | 0.90 |

**Figure S2:**
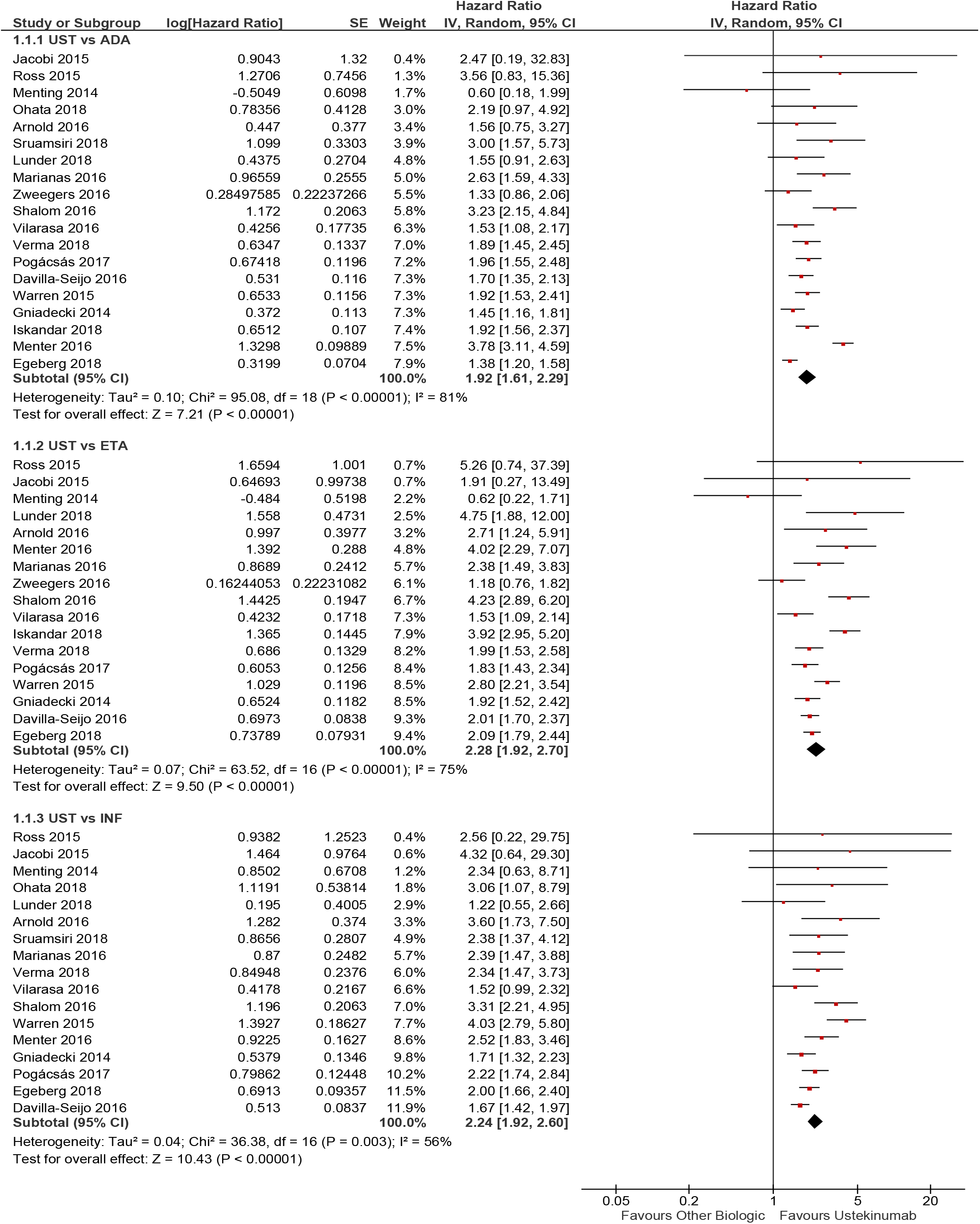
Comparative drug survival for ustekinumab vs. other biologics (adalimumab, etanercept, and infliximab) at 2-years.

**Figure S3:**
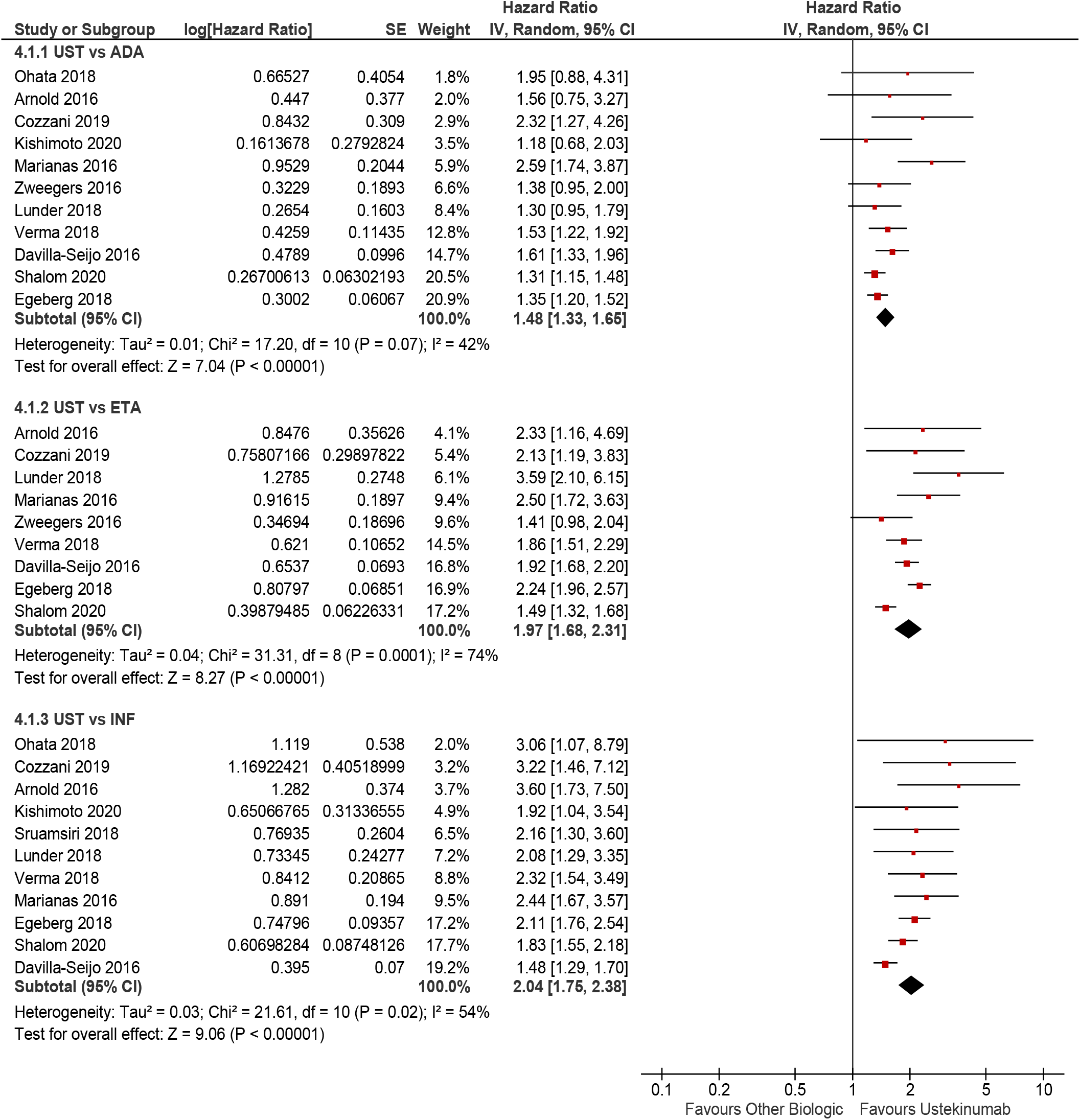
Comparative drug survival for ustekinumab vs. other biologics (adalimumab, etanercept, and infliximab) at 5 years.

**Figure S4:**
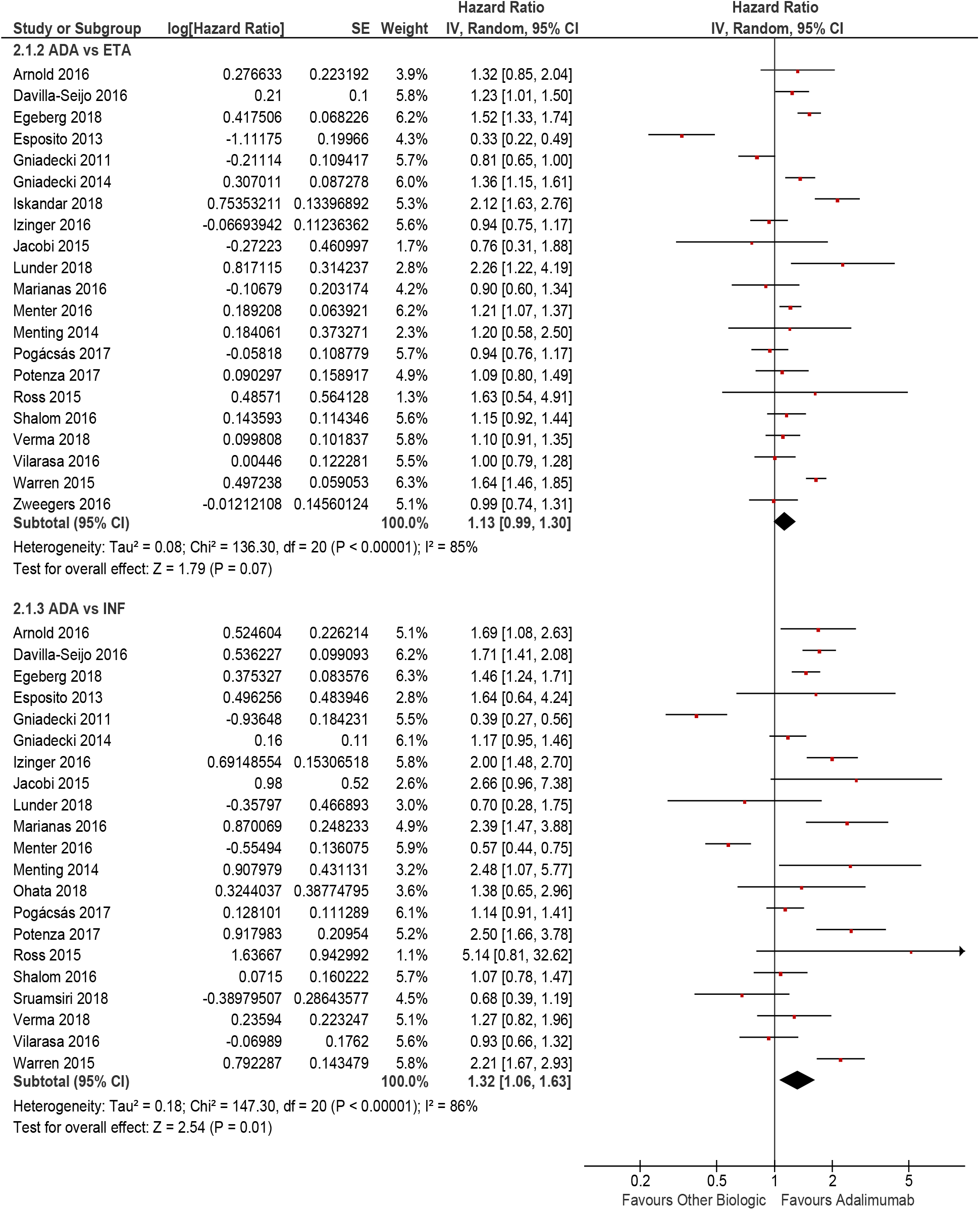
Comparative drug survival for adalimumab vs. other biologics (etanercept, and infliximab) at 2 years.

**Figure S5:**
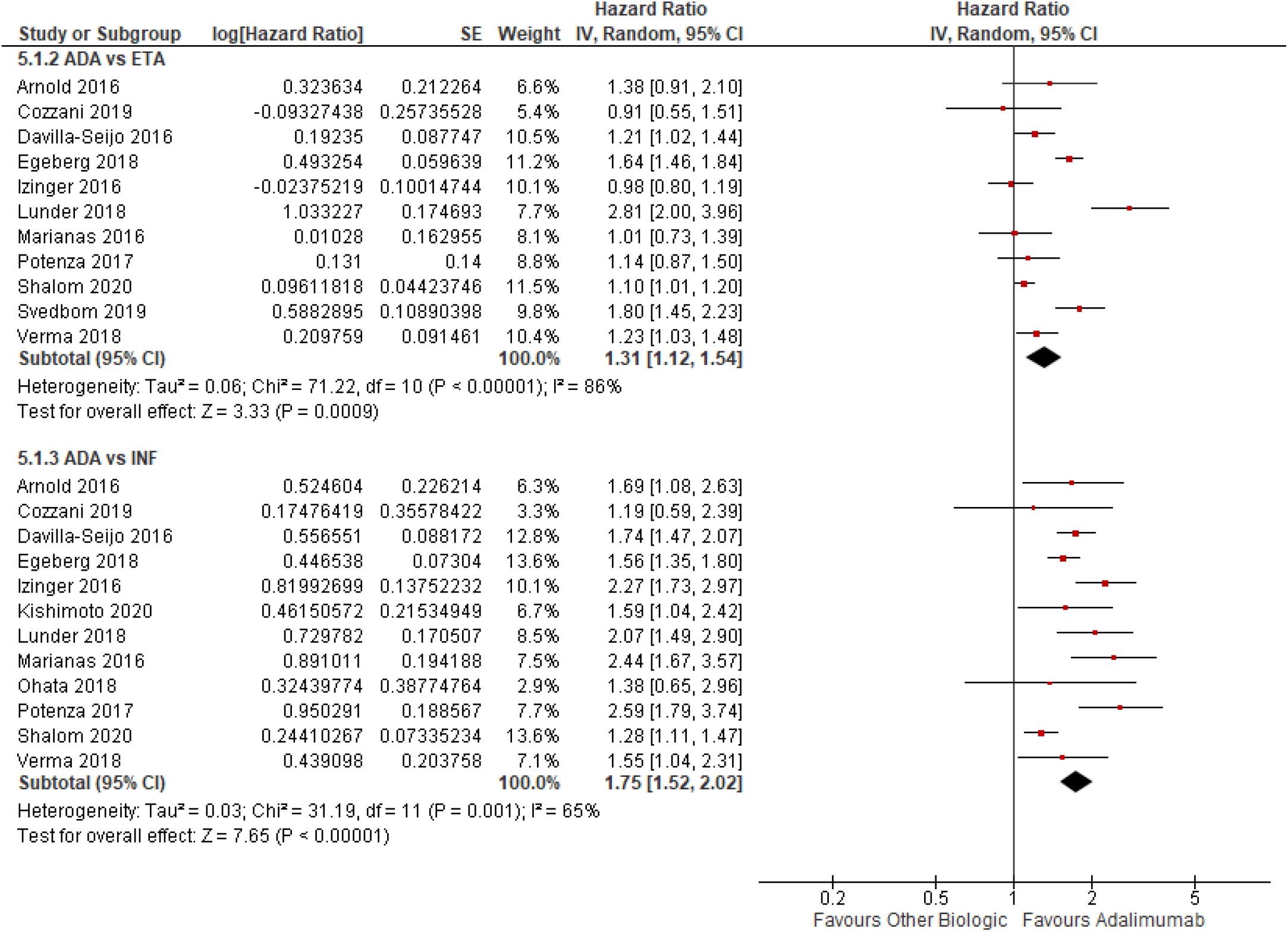
Comparative drug survival of adalimumab vs. other biologics (etanercept, and infliximab) at 5 years.

**Figure S6:**
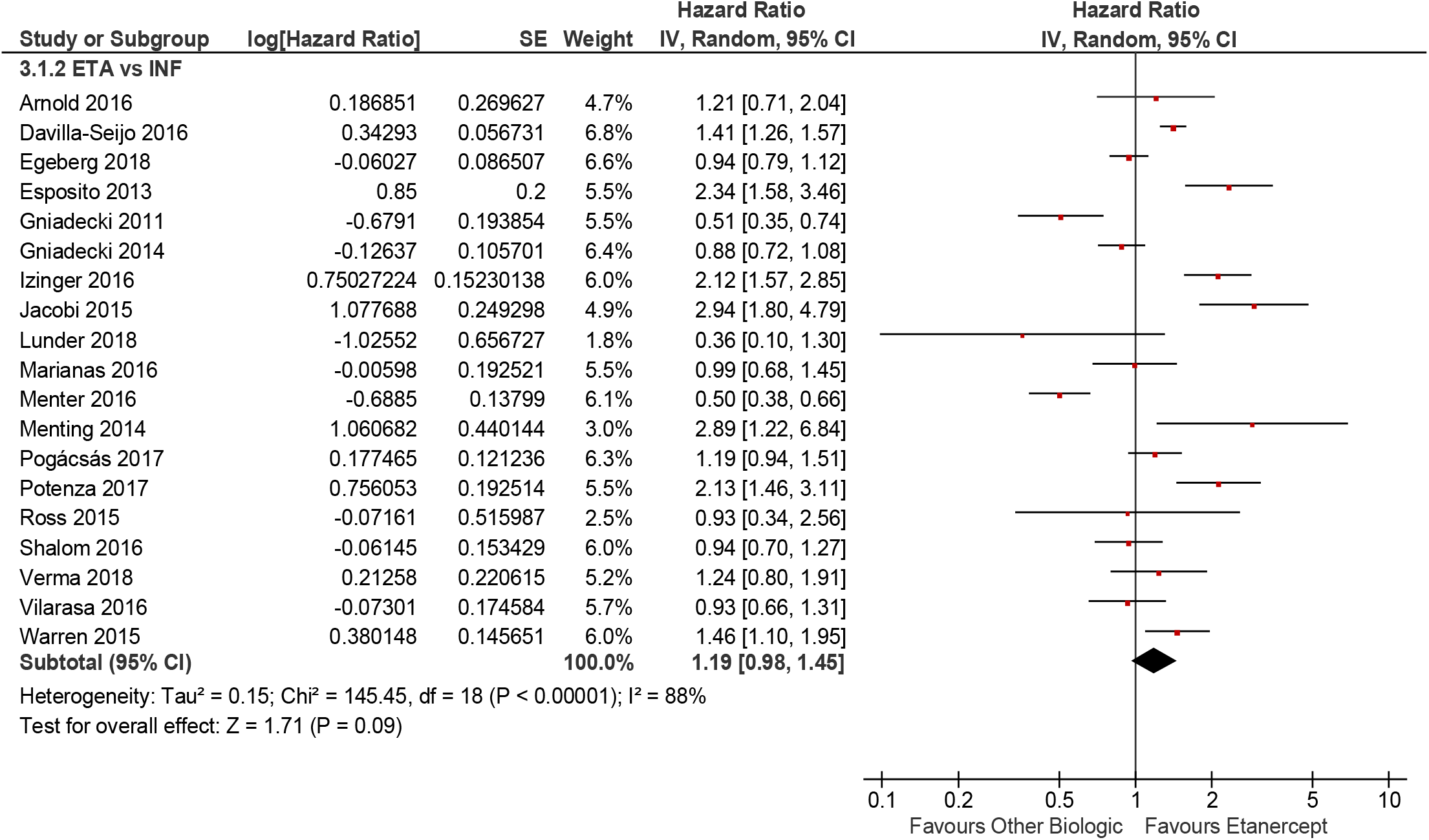
Comparative drug survival of etanercept vs. infliximab at 2 years.

**Figure S7:**
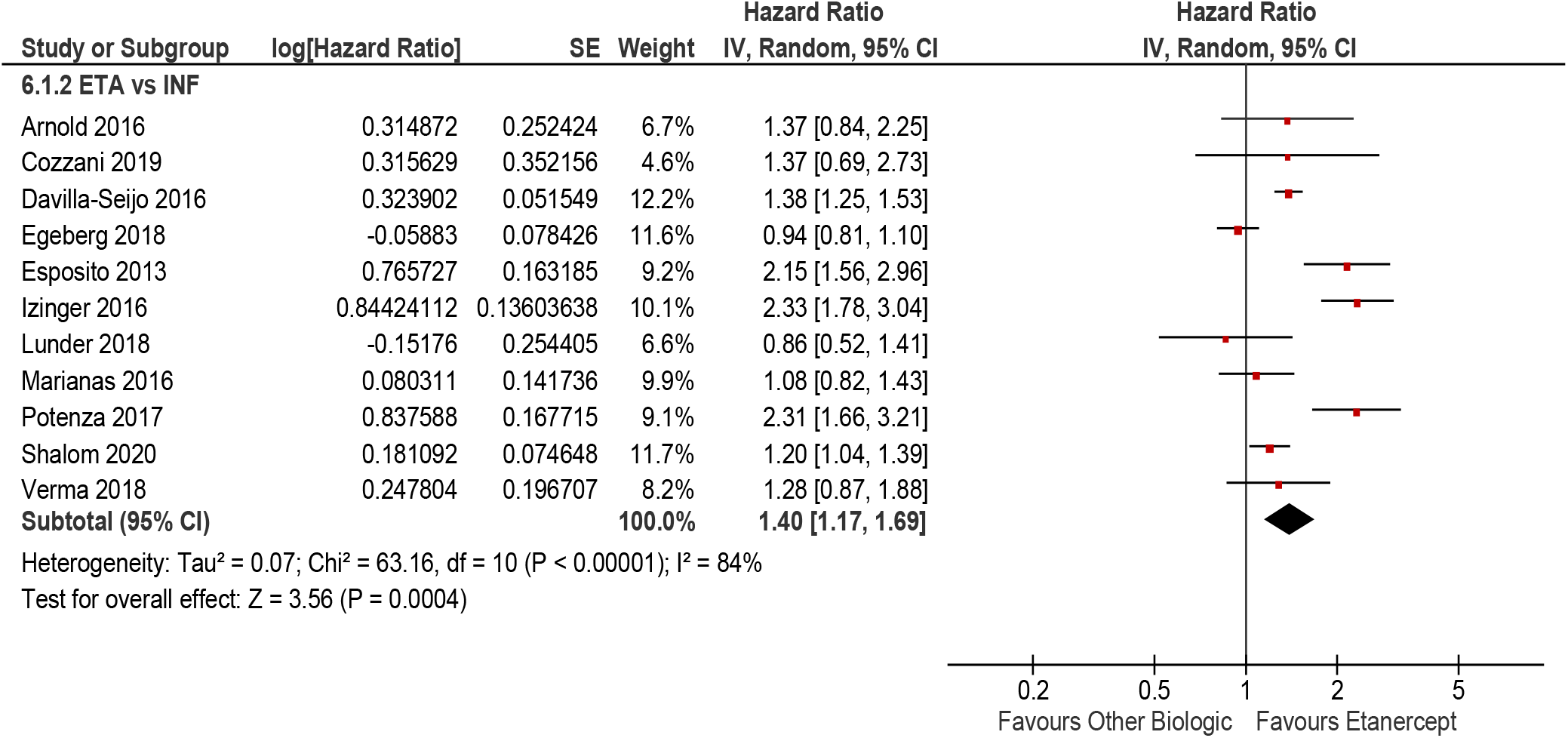
Comparative drug survival for etanercept vs. infliximab at 5 years.

**Figure S8:**
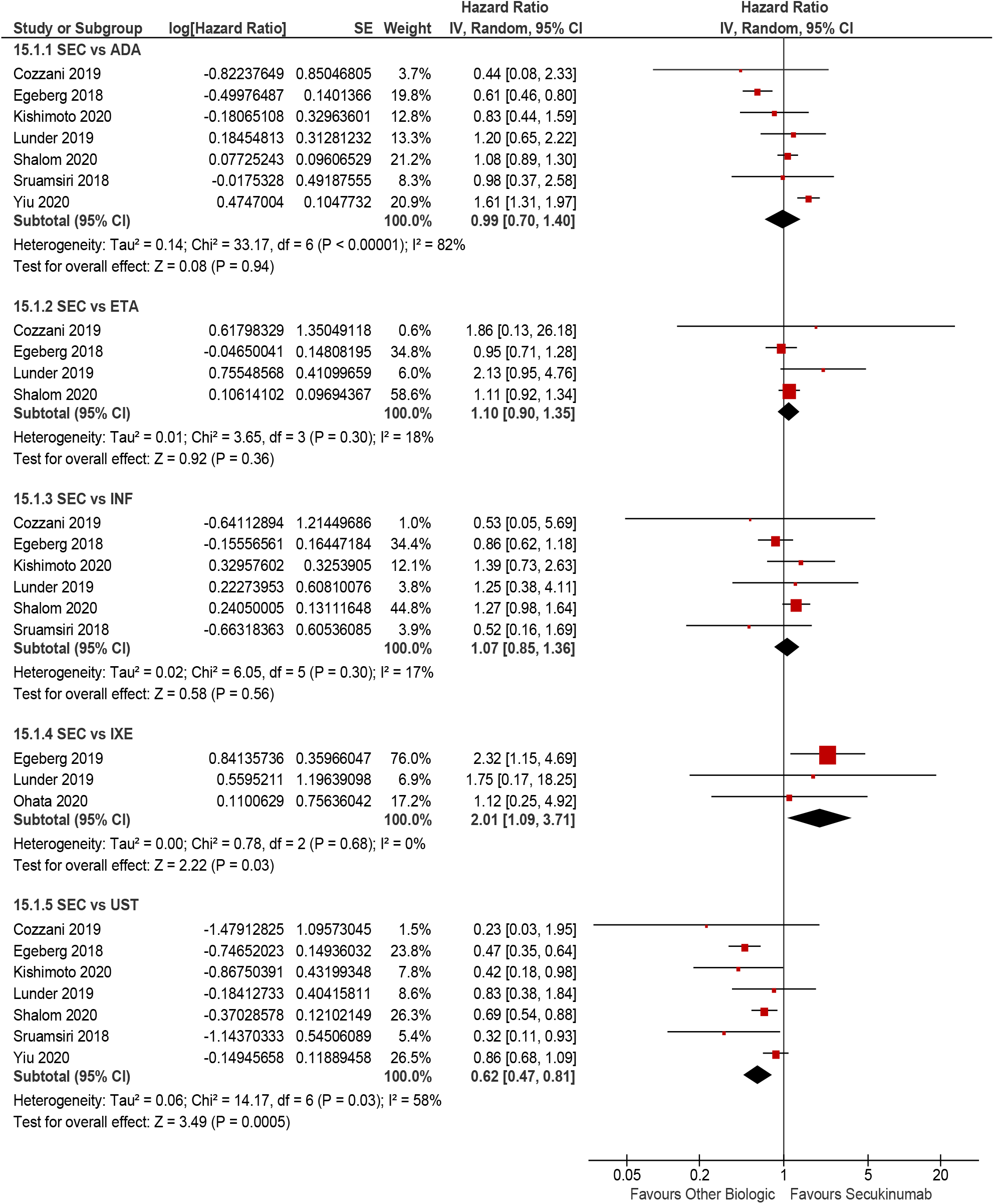
Comparative drug survival for secukinumab vs. other biologics (adalimumab, etanercept, infliximab, ixekizumab, and ustekinumab) at 1 year.

